# Mobile Life Skills Education Adoption Among Internally Displaced Persons in Northern Nigeria and Health Systems Implications for Equitable Mental Health Support: A cross-sectional study

**DOI:** 10.1101/2025.06.18.25329867

**Authors:** Andem Effiong Etim Duke, Ihoghosa Iyamu, Attila J Hertlendy, Bala I Harri, Abdulrahman Ibrahim, Abduljaleel Adejumo, Chisom Obi-Jeff, Sanni Yaya, Vincent Agyapong, Rita Orji, Ejemai Eboreime

**Author notes:** **Corresponding Author: Name:** Ejemai Eboreime, **Institution:** Faculty of Medicine, Department of Psychiatry, Dalhousie University, **Address:** Abbie J. Lane Building, 8th Floor, 5909 Veterans’ Memorial Lane, Halifax, NS B3H 2E2, Canada, **Email:**.

## Abstract

**Introduction:** Internally displaced persons (IDPs) face significant mental health challenges amidst severely disrupted health systems. Digital interventions offer promising pathways to deliver psychosocial support, yet critical gaps remain in understanding what determines their acceptability and adoption among vulnerable populations in conflict settings.

**Objective:** This study investigates the sociodemographic, psychosocial, technological, and cultural determinants of interest in mobile-based life skills education (mLSE) among IDPs in Nigeria, integrating four theoretical frameworks to generate actionable insights for equitable digital mental health service delivery in fragile settings.

**Methods:** We analyzed cross-sectional data from 220 IDPs in the Durumi and Wassa camps of Abuja, Nigeria. Variable selection employed elastic net regression, identifying 22 key predictors. Modified robust Poisson regression estimated prevalence ratios for mLSE interest, with interaction effects modeled to capture demographic intersectionalities.

**Results:** Among participants, 48.6% expressed interest in mLSE, with significant disparities across age, education, and camp location. Young adults aged 20-24 with prior counseling experience showed substantially higher interest (APR=3.49, 95% CI 1.72-7.10), while males with counseling history demonstrated markedly lower engagement (APR=0.33, 95% CI 0.19-0.57). Secondary education strongly predicted interest (APR=2.27, 95% CI 1.59-3.26), as did residence in the Wassa camp (APR=1.64, 95% CI 1.21-2.23). Notably, males aged 30-34 exhibited minimal interest (APR=0.09, 95% CI 0.01-0.75), revealing critical gender-age intersections.

**Conclusions:** These findings reveal actionable patterns for strengthening digital mental health service delivery in displacement settings. Health systems in fragile contexts must develop digitally delivered interventions that are culturally responsive, gender-sensitive, and age-appropriate, while addressing educational and technological barriers. Leveraging prior service engagement appears critical for sustainable implementation. This study provides a roadmap for policymakers and implementers to design equitable digital mental health interventions that address the disparate needs of displaced populations.

## Introduction

Internally displaced persons (IDPs) who are forced to leave their homes due to conflict, natural disasters, or crises but remain within their country’s borders face unique mental health challenges exacerbated by limited access to healthcare and social stigma [1,2,3,4]. With more than 75.9 million IDPs globally, particularly in regions like Sub-Saharan Africa, the Middle East, and Eastern Europe, there is a pressing need for scalable, accessible mental health interventions [5]. Recent crises, such as those in Ukraine and Sudan, have amplified the urgency of addressing mental health needs in displacement contexts [5,6,7,8,9]

Among the key challenges IDPs face are the disruption of social support systems and the limited availability of mental health services [10,11]. Life skills education (LSE) has shown promise in enhancing resilience and improving mental health among displaced populations [10,11,12,13,14,15,16]. Mobile technology presents a promising solution to deliver LSE, especially in settings with limited or disrupted traditional services. For example, mobile-based interventions in Mozambique and Ethiopia have proven effective in reaching displaced populations where traditional delivery mechanisms are compromised [17,18,19].

The effectiveness of mobile-based LSE (mLSE) is well-documented in randomized controlled trials, which have shown measurable improvements in psychological wellbeing and coping mechanisms among participants [20,21]. Digital platforms, such as the mobile app BetterMe: Mental Health, which combines guided meditations, Cognitive Behavioral Therapy (CBT), and mindfulness exercises, are increasingly used to address mental health needs in crisis settings [22,23,24]. These digital interventions have demonstrated success in Ukraine, where over 3.6 million people remain internally displaced as of October 2024 [25]. Initiatives like U-RISE (Ukraine’s displaced people in the European Union (EU): Reach out, Implement, Scale-up and Evaluate interventions promoting mental wellbeing) and BetterMe, developed in collaboration with the World Health Organization (WHO) and United Nations International Children’s Education Fund (UNICEF), provide evidence-based psychosocial support and offer valuable lessons for implementing similar strategies in African contexts [26,27,28].

This study builds on four complementary frameworks to examine the factors influencing the adoption of mLSE among IDPs in Abuja, Nigeria. The Socio-Ecological Model (SEM) posits that multiple interconnected layers of influence shape human behavior: individual factors (e.g., personal beliefs and skills), interpersonal connections (e.g., family and friends), community influences (e.g., cultural norms), organizational structures (e.g., institutional rules), and policy environment (e.g., laws and regulations) [29]. The Diathesis-Stress Theory (DST) explains how displacement amplifies psychological vulnerabilities [30], while the Technology Acceptance Model (TAM) examines the role of digital literacy in the adoption of mLSE [31]. Finally, the Cultural Influence on Mental Health (CIMH) framework emphasizes the need for culturally sensitive interventions to enhance engagement and effectiveness [2]. By applying these frameworks, the study aims to inform the design of accessible, culturally relevant, and scalable mental health support programs for IDPs in conflict-affected settings.

Specifically, this study addresses the following research questions: Which sociodemographic, psychosocial, technological, and cultural factors are associated with interest in adopting mLSE among IDPs in Durumi and Wassa camps, and what do these patterns imply for equitable mental health service delivery in fragile settings? To what extent do interactions among psychological counseling history, age, gender, and education shape IDPs’ interest in mLSE interventions, and how can health and social care systems leverage these insights to tailor digital psychosocial support?

## Methods

This study adhered to the Strengthening the Reporting of Observational Studies in Epidemiology (STROBE) guidelines for cross-sectional studies. In addition, this study presents a cross-sectional analysis of baseline (pre-intervention) survey data collected by our research team from September 2024 to January 2025 as part of the ongoing Rebuilding Emotional Stability and Strength Through Therapeutic and Life-Skills Education for Internally Displaced Persons in Nigeria (RESETTLE-IDPs) trial <u>(NCT06412679)</u> [12,33]. RESETTLE-IDPs is a hybrid type II effectiveness-implementation cluster randomized controlled trial evaluating the clinical effectiveness and implementation outcomes of a contextually adapted life skills education (LSE) program among internally displaced persons (IDPs) in Northern Nigeria [12,33]. The RESETTLE-IDPs trial aims to strengthen psychosocial resilience, enhance coping strategies, and facilitate the sustainable integration of IDPs into host communities through structured psychosocial interventions [12,33].

Survey respondents were recruited from two IDP camps in Durumi and Wassa, Abuja, Federal Capital Territory (FCT), North Central Nigeria. Although these sites are sometimes called ‘settlements’ due to their lack of formal infrastructure, the terms ‘camps’ and ‘settlements’ are often used interchangeably in humanitarian contexts. Similarly, this study uses both terms interchangeably to refer to these locations. The analytical sample for this cross-sectional study consisted of 220 respondents. Informed consent or assent (for minors under 18 years of age) was obtained from all participants.

The outcome was interest in mLSE. mLSE encompasses structured educational content delivered via mobile phone technologies designed to equip displaced persons with psychosocial skills, including emotional regulation, effective communication, problem-solving, and stress management. These skills are intended to assist IDPs in coping effectively with the adverse impacts of displacement, enhance their resilience, and improve their overall mental wellbeing.

In the survey, this interest was referred to as “mobile learning potential” and defined as the respondent’s interest in using their mobile phone to receive education or training on how to cope with life challenges. Initially, the responses were categorized as “yes,” “no,” or “unsure.” For analysis, we dichotomized interest in mLSE into ‘yes’ (indicating interest, coded as 1) and ‘no’ (including ‘unsure’ responses, coded as 0).

Sociodemographic factors were assessed as covariates. These included the participant’s age, gender, education, employment status, and state of residence before displacement. Other independent variables were psychological counseling history and duration of stay in the IDP camp or settlement.

In this study, we use the term gender to denote the categories male and female as reported by participants, which correspond to the sex assigned at birth. Although gender broadly encompasses socially constructed roles, identities, and behaviors, our survey collected only male and female categories. We recognize that biological sex categorizations include individuals who are intersex or have differences of sex development (DSD), and that our binary framework does not capture these variations or other gender-diverse identities.

All statistical analyses were conducted using R version 4.4.2 (The R Foundation for Statistical Computing, Vienna, Austria) and Stata 16 (StataCorp LLC, College Station, Texas, United States).

Categorical variables were summarized as frequencies and percentages. Distributions of participant characteristics by interest in mLSE were explored using chi-square (χ²) tests as an initial descriptive step (Table 1).

**Table 1:** Distribution of Study Characteristics by Life Skills Education, n=220 IDPs.

| Variable | Category | n | % | $\chi^2$ | p-value |
| --- | --- | --- | --- | --- | --- |
| AGE | 13-19 years | 14 | 6.36 | 49.5 | <0.0001 |
|  | 20-24 years | 28 | 12.73 |  |  |
|  | 25-29 years | 47 | 21.36 |  |  |
|  | 30-34 years | 57 | 25.91 |  |  |
|  | 35+ years | 74 | 33.64 |  |  |
| GENDER | Female | 173 | 78.64 | 1.86 | 0.17 |
|  | Male | 47 | 21.36 |  |  |
| EDUCATION | Islamic education | 54 | 24.55 | 91.41 | <0.0001 |
|  | No formal education | 73 | 33.18 |  |  |
|  | Primary education | 39 | 17.73 |  |  |
|  | Secondary education or Higher | 54 | 24.55 |  |  |
| EMPLOYMENT | No | 165 | 75 | 2.68 | 0.10 |
|  | Yes | 55 | 25 |  |  |
| **BARRIER** | Accessibility/Affordability | 84 | 38.18 | 49.5 | <0.0001 |
|  | None | 20 | 9.09 |  |  |
|  | Technological | 116 | 52.73 |  |  |
| **IDP CAMP LOCATION** | Durumi IDP Camp | 146 | 66.36 | 7.68 | 0.021 |
|  | Wassa IDP Camp | 74 | 33.64 |  |  |
| **ENGLISH** | No | 186 | 84.55 | 12.47 | <0.0001 |
|  | Yes | 134 | 15.45 |  |  |
| **HAUSA** | No | 49 | 22.27 | 1.06 | 0.30 |
|  | Yes | 171 | 77.73 |  |  |
| **MOBILE PHONE USE** | Calls | 153 | 69.55 | 39.39 | <0.0001 |
|  | Miscellaneous | 67 | 30.45 |  |  |
| **NO SOCIAL MEDIA PLATFORM** | False | 35 | 15.91 | 43.96 | <0.0001 |
|  | True | 185 | 84.09 |  |  |
| **PSYCHOLOGICAL COUNSELING HISTORY** | False | 139 | 63.18 | 8.80 | 0.0030 |
|  | True | 81 | 36.82 |  |  |
| **DURATION IN CAMP** | 0-5 years | 146 | 66.36 | 47.04 | <0.0001 |
|  | 6+ years | 74 | 33.64 |  |  |

A correlation matrix was generated to assess multicollinearity among the covariates, and variance inflation factor (VIF) analysis was conducted. Due to the presence of highly correlated predictors, elastic net regression was employed for variable selection. This method combines the penalty functions of ridge regression, which minimizes the impact of collinearity by shrinking coefficients, and the least absolute shrinkage and selection operator (LASSO), which facilitates variable selection by setting some coefficients to zero. By combining these techniques, elastic net regularization is particularly effective in identifying relevant predictors when multicollinearity is present.

The elastic net model identified 22 predictors associated with interest in mLSE (Figure 1). The most influential factors included a lack of access to social media platforms, being 35 years or older, having a secondary education or higher, and a duration of displacement exceeding six years. Other selected predictors included technological barriers, age 20 to 24 years, language spoken, and settlement location. Model performance was assessed using the area under the receiver operating characteristic curve (AUC ROC), which indicated high predictive accuracy (AUC = 0.89; Figure 2).

**Figure 1:**
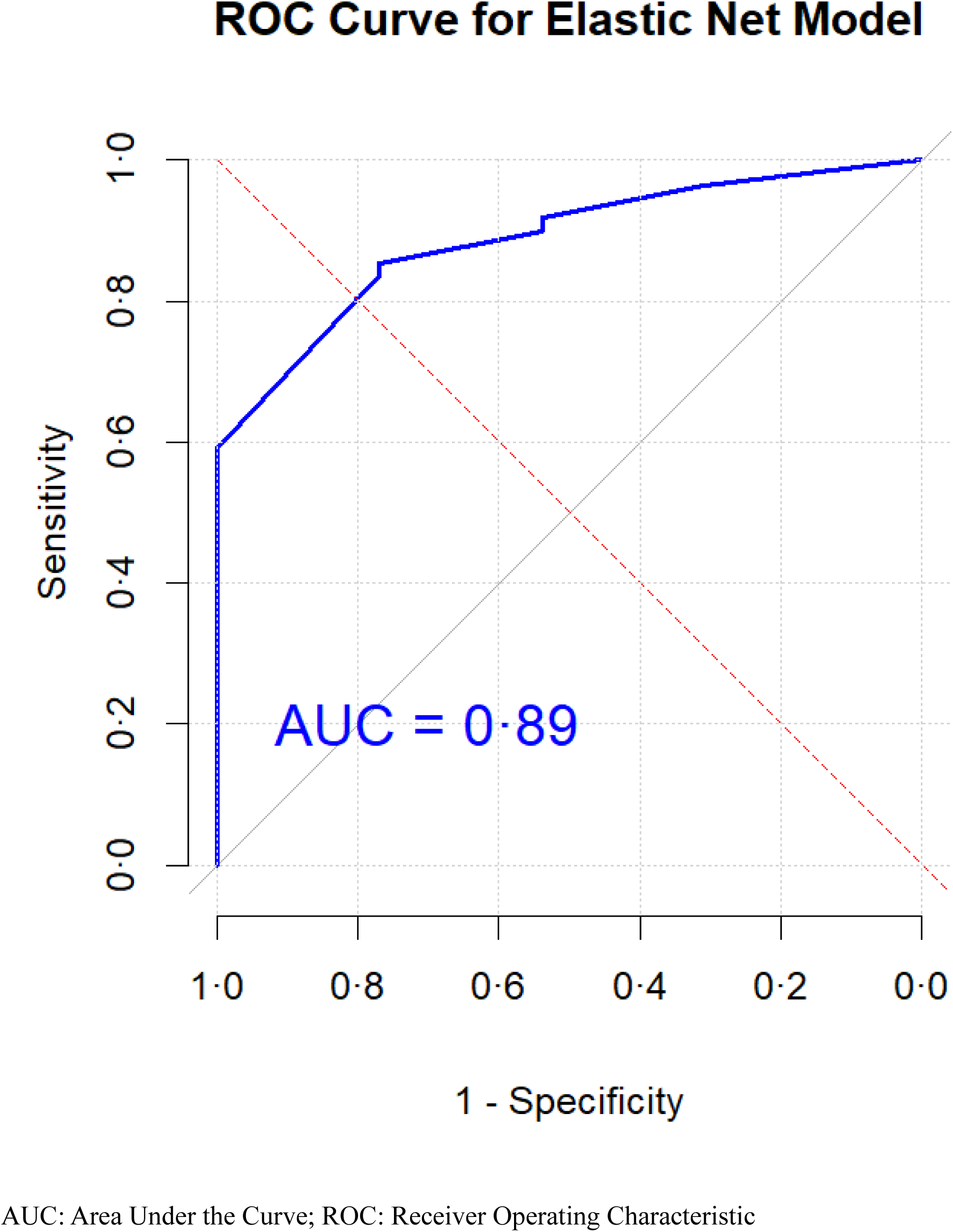
AUC and ROC analysis for Elastic Net Model.

**Figure 2:**
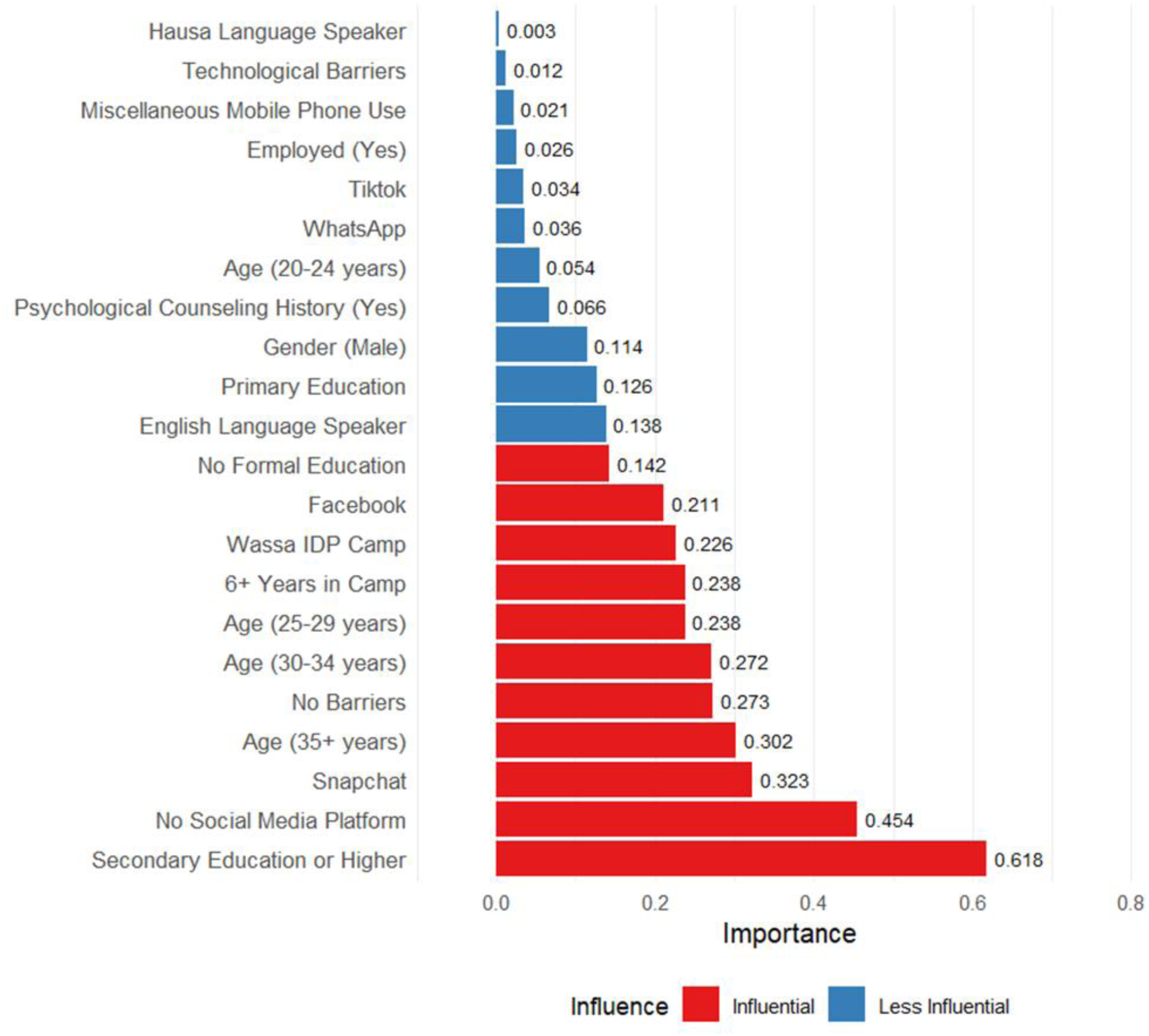
Variable importance plot of mLSE for IDPs.

Following elastic net selection, high correlations remained among some retained variables (Figure 3). To address this and estimate covariate-adjusted prevalence ratios (PRs) directly, a modified robust Poisson regression model was utilized. This method is particularly suitable for binary outcomes in cross-sectional studies and offers a more interpretable alternative to logistic regression. The robust Poisson approach employs a sandwich variance estimator, which accounts for potential dependence in the data, making it suitable in contexts where correlated outcomes are likely to occur.

**Figure 3:**
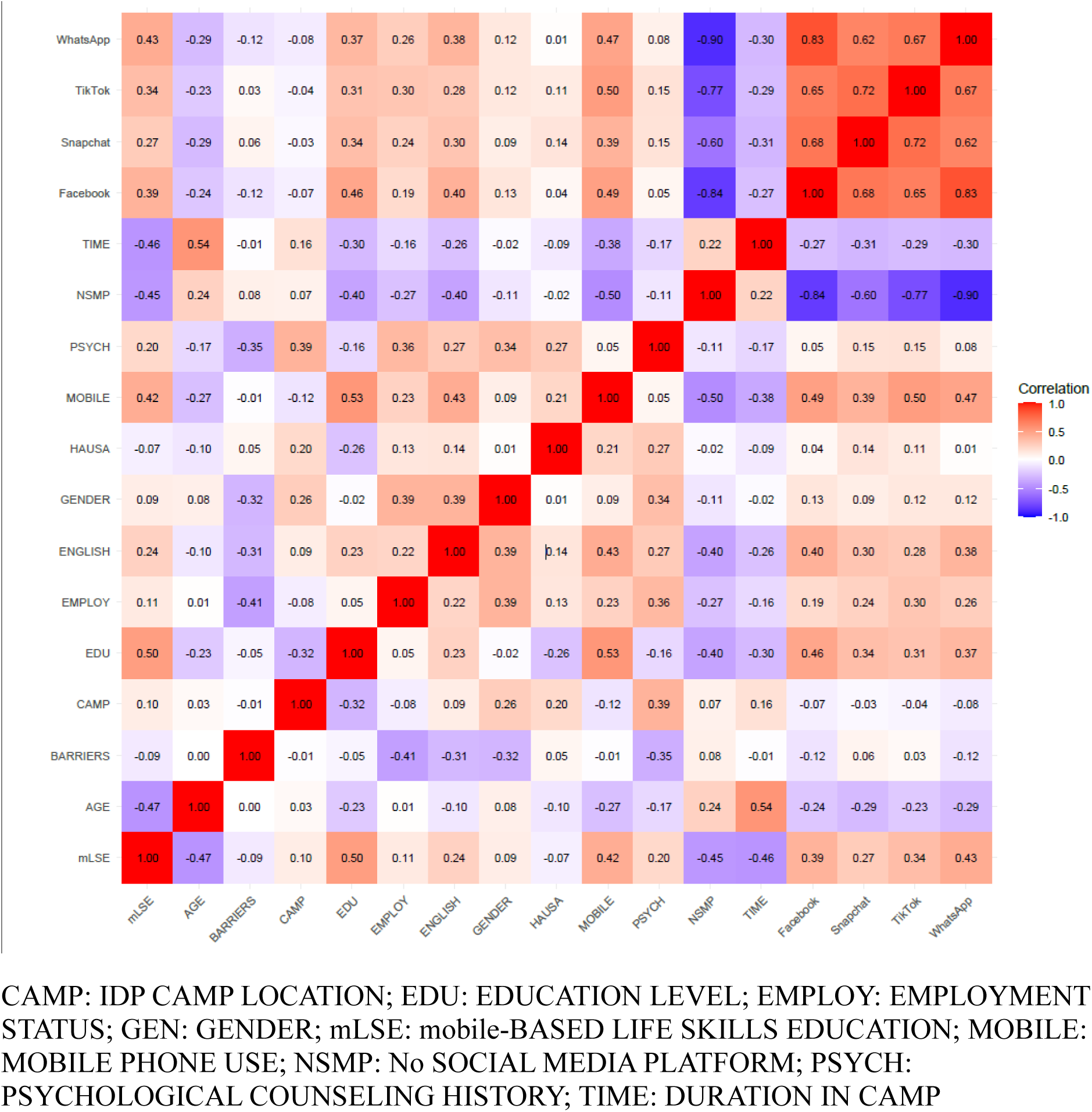
Heatmap and pairwise correlation matrix depicting relationships between variables and mLSE.

The predictors identified by the elastic net regression were entered into the robust Poisson model to estimate adjusted prevalence ratios (APRs) and 95% confidence intervals (CIs) (Table 2 and Table A.1, Figure 4).

**Figure 4:**
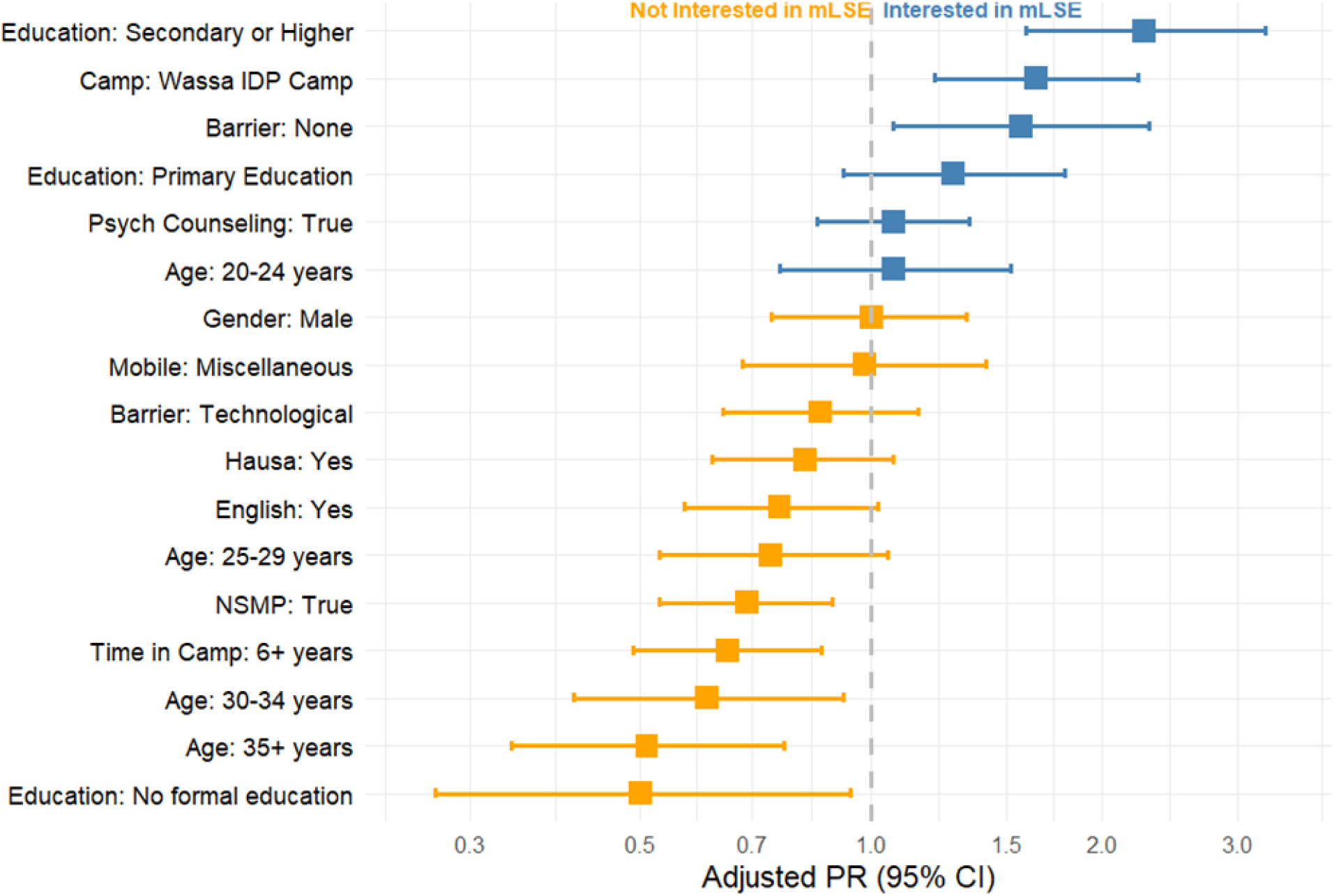
Adjusted Prevalence Ratio (PR) of IDP Characteristics and Interest in mLSE.

**Table 2.** Adjusted Prevalence Ratios for interest in mLSE.

| Variable | Category | APR | Robust SE | z | P>\|z\| | 95% CI |
| --- | --- | --- | --- | --- | --- | --- |
| AGE | 13-19 years | 1.00 (base) | - | - | - | - |
|  | 20-24 years | 1.07 | 0.19 | 0.38 | 0.70 | 0.76-1.52 |
|  | 25-29 years | 0.74 | 0.13 | -1.69 | 0.090 | 0.53-1.05 |
|  | 30-34 years | 0.61 | 0.13 | -2.37 | 0.018 | 0.41-0.92 |
|  | 35+ years | 0.51 | 0.11 | -3.19 | 0.001 | 0.34-0.77 |
| GENDER | Female | 1.00 (base) | - | - | - | - |
|  | Male | 1.00 | 0.15 | -0.03 | 0.97 | 0.74-1.33 |
| EDUCATION | Islamic education | 1.00 (base) | - | - | - | - |
|  | No formal education | 0.50 | 0.16 | -2.14 | 0.032 | 0.27-0.94 |
|  | Primary education | 1.28 | 0.22 | 1.47 | 0.14 | 0.92-1.79 |
|  | Secondary education or Higher | 2.27 | 0.42 | 4.48 | <0.0001 | 1.59-3.26 |
| EMPLOYMENT | No | 1.00 (base) | - | - | - | - |
|  | Yes | 0.98 | 0.17 | -0.11 | 0.91 | 0.67-1.38 |
| BARRIER | Accessibility/Affordability | 1.00 (base) | - | - | - | - |
|  | None | 1.57 | 0.30 | 2.32 | 0.020 | 1.07-2.30 |
|  | Technological | 0.86 | 0.13 | -1.04 | 0.30 | 0.64-1.15 |
| IDP CAMP LOCATION | Durumi IDP Camp | 1.00 (base) | - | - | - | - |
|  | Wassa IDP Camp | 1.64 | 0.26 | 3.19 | 0.001 | 1.21-2.23 |
| ENGLISH | No | 1.00 (base) | - | - | - | - |
|  | Yes | 0.76 | 0.11 | -1.84 | 0.066 | 0.57-1.02 |
| HAUSA | No | 1.00 (base) | - | - | - | - |
|  | Yes | 0.82 | 0.11 | -1.47 | 0.14 | 0.62-1.07 |
| MOBILE PHONE USE | Calls | 1.00 (base) | - | - | - | - |
|  | Miscellaneous | 0.98 | 0.18 | -0.12 | 0.90 | 0.68-1.41 |
| NO SOCIAL MEDIA PLATFORM | False | 1.00 (base) | - | - | - | - |
|  | True | 0.69 | 0.09 | -2.84 | 0.004 | 0.53-0.89 |
| PSYCHOLOGICAL COUNSELING HISTORY | False | 1.00 (base) | - | - | - | - |
|  | True | 1.07 | 0.12 | 0.59 | 0.56 | 0.85-1.34 |
| DURATION IN CAMP | 0-5 years | 1.00 (base) | - | - | - | - |
|  | 6+ years | 0.65 | 0.09 | -2.99 | 0.003 | 0.49-0.86 |

The final model retained variables not selected by the elastic net but theoretically grounded in the study’s guiding frameworks and directly aligned with the research questions, including psychological counseling history and gender.

Interaction terms were specified to examine whether the association between psychological counseling history and mLSE interest varied across age, gender, and education. Marginal standardized probabilities were computed to facilitate the interpretation of these interactions, and the results were visualized using predictive margin plots (Table A.1; Figures A.1-A.4). Marginal standardization is a suitable method for inferring population-level effects.

E-values were calculated for all key estimates to assess the robustness of the observed associations to unmeasured confounding (Table A.3).

## Results

Descriptive and baseline characteristics are presented in Table 1.

Among the 220 IDPs, 107 (48.64%) reported interest in mLSE. Age (χ² = 49.50, p <0.0001) and education level (χ² = 91.41, p < 0.0001) were associated with interest, with younger IDPs and those with higher education levels being more likely to show interest. Camp location (χ² = 7.68, p = 0.021), English language (χ² = 12.47, p < 0.0001), and mobile phone use (χ² = 39.39, p < 0.0001) were also found to be associated with interest.

Significant associations were observed between interest in mLSE and barriers to technological or financial accessibility (χ² = 49.50, p < 0.001), psychological counseling history (χ² = 8.80, p = 0.0030), social media usage (χ² = 43.96, p < 0.001), and duration of stay in the camp (χ² = 47.04, p < 0.001). No significant associations were observed for gender, employment status, or primary language use.

We observed a lower adjusted prevalence of interest in mLSE among older age groups compared to the reference group aged 13-19. The adjusted prevalence ratio for interest was 0.61 (95% CI, 0.41-0.92; p = 0.018) for those aged 30-34 years vs 13-19 years, and 0.51 (95% CI, 0.34-0.77; p = 0.001) for those aged 35 years or older vs 13-19 years (Table 2, Figure 4).

Higher education levels were associated with a greater adjusted prevalence of interest in mLSE than those with Islamic education. IDPs with secondary education or higher had an APR of 2.27 (95% CI, 1.59-3.26; p <0.0001), while those with no formal education had a reduced APR of 0.50 (95% CI, 0.27-0.94; p = 0.032) (Table 2, Figure 4).

IDPs residing in the Wassa IDP Camp had a higher adjusted prevalence of interest in mLSE than those in the Durumi IDP Camp (APR, 1.64; 95% CI, 1.21-2.23; p = 0.0010). IDPs experiencing neither accessibility/affordability nor technological barriers showed a greater adjusted prevalence of interest (APR, 1.57; 95% CI, 1.07-2.30; p = 0.020) compared to those experiencing accessibility/affordability barriers alone.

Non-social media users had a lower adjusted prevalence of interest in mLSE (APR, 0.69; 95% CI, 0.53-0.89; p = 0.0040) than social media users. Furthermore, IDPs with longer durations in camp (6+ years) were less interested in mLSE (APR, 0.65; 95% CI, 0.49-0.86; p = 0.0030) compared to those with shorter stays (0-5 years) (Table 2, Figure 4).

The adjusted prevalence of interest in MLSE was slightly higher among IDPs with a history of psychological counseling (APR 1.07; 95% CI 0.85–1.34; p = 0.56) compared to those without counseling. However, this difference was not statistically significant (Table 2, Figure 4).

Interaction effect estimates pointed to conditional associations between age groups, gender, education level, psychological counseling history, and interest in mLSE. For male IDPs aged 20-24 years, the prevalence of interest in mLSE was higher (APR, 1.75; 95% CI, 1.06-2.88; p = 0.028) compared to females in the reference group (13-19 years). Conversely, males aged 30-34 exhibited a lower prevalence of interest (APR, 0.09; 95% CI, 0.01-0.75; p = 0.026), suggesting a substantial reduction in interest for this subgroup.

IDPs aged 20-24 years with a history of psychological counseling had a higher prevalence of interest in mLSE (APR, 3.49; 95% CI, 1.72-7.10; p = 0.0010). In contrast, males with a history of psychological counseling since displacement showed a reduced prevalence of interest in mLSE (APR, 0.33; 95% CI, 0.19-0·57; p < 0.0001).

For individuals with primary education and a history of psychological counseling since displacement, the prevalence of interest in mLSE also increased (APR, 2.76; 95% CI, 1.49-5.10; p = 0.001), highlighting a significant positive association between primary education level, psychological counseling history, and interest in mLSE.

Marginal standardization indicated nuanced interactions and distinct patterns across subgroups. Predicted probabilities further illustrated the complex influence of age, gender, education, and psychological counseling history on interest in mLSE (Figs. A.1-A.4).

Sensitivity analysis showed relatively modest E-values (Table A.3).

## Discussion

Guided by four theoretical frameworks, this cross-sectional study examined sociodemographic, psychosocial, technological, and cultural determinants and their interactions of interest in mLSE among IDPs in the Durumi and Wassa camps of Abuja, Northern Nigeria, and evaluated implications for strengthening equitable digital mental health service delivery within fragile health systems. Collectively, these factors provide actionable insights for developing and implementing tailored mLSE interventions in IDP settings.

A key finding was the association between age and interest in mLSE. Younger IDPs, especially those aged 20 to 24, showed the highest interest levels, while older groups, notably those aged 30 to 34 and 35 and above, exhibited reduced engagement. This trend is consistent with the TAM, which posits that individuals are more likely to adopt technologies they perceive as easy to use and valuable [31]. Younger participants may have greater exposure to mobile platforms and more positive attitudes toward digital learning. In contrast, older individuals may lack digital literacy or perceive mLSE as less relevant to daily challenges. Addressing these disparities requires user-friendly interfaces, digital training, and content that demonstrates practical relevance for older IDPs.

Educational attainment also played a crucial role in the interest in mLSE. IDPs with secondary education or higher showed significantly more interest in mLSE compared to those with no formal or Islamic education. This supports prior findings that education enhances digital navigation skills and receptivity to new learning modalities [29]. The association emphasizes the importance of designing inclusive mLSE content that utilizes simplified language, visual aids, and peer support to accommodate those with limited literacy.

A history of psychological counseling was found to interact with several demographic factors. Individuals with prior counseling experience, particularly younger adults, were more inclined toward mLSE, perhaps due to increased mental health literacy and familiarity with structured support. Nonetheless, this pattern diverged among males, where a history of counseling was associated with reduced interest. This finding highlights how gender norms and expectations may influence help-seeking behavior and acceptance of digital interventions. In line with the CIMH model [34], our findings suggest that male IDPs may be more affected by stigma or expectations of self-reliance. Incorporating culturally relevant messaging, male role models, and gender-sensitive outreach could improve uptake among this group [35,36].

The community context also shaped mLSE interest. IDPs in the Wassa camp expressed higher interest than those in Durumi, indicating the potential influence of local infrastructure, communal norms, and prior exposure to health programs. Similarly, those displaced for more than six years were less interested in mLSE than those with shorter displacement durations, possibly due to the development of entrenched coping routines or disillusionment with digital initiatives. These findings reflect the SEM [29], which emphasizes that behavior is shaped by factors across multiple levels, including community and organizational dynamics.

Technological access was another key determinant. IDPs who reported technological or affordability barriers were less interested in mLSE. Those without access to social media platforms had lower engagement levels. This digital divide highlights infrastructural inequalities within IDP camps that must be addressed through targeted policy interventions, including the provision of subsidized mobile devices, enhanced connectivity, and offline-compatible mLSE applications.

Comparative insights from other LMIC displacement contexts reinforce these findings [37]. For example, studies in Bangladesh and among Rohingya refugees have shown that education levels, cultural attitudes, and prior mental health service exposure shape receptiveness to digital tools [38,39]. Nonetheless, the relatively higher uptake of mLSE among Nigerian IDPs with counseling experience may reflect the dynamics in urban displacement settings, such as Abuja, where informal support networks and access to information are stronger [40,41,42].

Cultural considerations remain crucial [43,44]. Our findings suggest that cultural beliefs influence the recognition and management of psychological distress [43, 44, 45,46]. For example, the lower interest among males aged 30 to 34 may indicate reluctance to engage in mental health services [46,47]. Counseling history was a positive predictor of mLSE interest across most groups, suggesting that exposure to structured formal care fosters openness to digital interventions. To increase acceptability, mLSE programs must be embedded within culturally familiar practices, integrating local idioms, healing narratives, and communal support systems.

From an implementation perspective, the study offers practical guidance. Programs should integrate digital literacy modules tailored to varying age and education levels. Culturally sensitive and gender-specific strategies are crucial for reaching populations that may face stigma or lack confidence in using digital tools. Leveraging existing social media usage can serve as a gateway to engage users while also addressing access issues through infrastructure support.

Building trust through community leaders and facilitators can increase the credibility and relevance of mLSE content [48]. Aligning mLSE delivery with camp-based psychosocial initiatives can support seamless integration and sustainability.

The study’s alignment with the DST reinforces the importance of psychological history in shaping receptivity to interventions [27]. Individuals with prior counseling likely have a higher awareness of emotional challenges and more readiness to engage with mental health resources. Recognizing this readiness can guide targeting and stratification strategies in IDP settings mLSE rollout.

Limitations include the cross-sectional design, which precludes our ability to make some causal inferences. The study’s geographic focus on Abuja may limit generalizability, and self-reported data may introduce biases regarding mental health and technology use. Although robust statistical methods were employed, residual confounding cannot be ruled out entirely. Future research should utilize longitudinal and mixed-methods designs to assess mLSE uptake and investigate how mLSE can complement traditional support mechanisms.

Interest in mLSE among Nigerian IDPs is influenced by modifiable factors, including age, education, gender, and psychological history, all of which are situated within cultural and structural contexts. Young adults emerged as a particularly receptive group, while older individuals and men require targeted engagement strategies. Cultural beliefs, access to technology, and community dynamics significantly influence receptiveness. When designed with demographic sensitivity and cultural awareness, mobile-based interventions such as mLSE can offer scalable support for mental health in displacement settings. These findings guide the development of equitable, accessible, and sustainable digital solutions that integrate the lived experiences of displaced populations and strengthen service delivery.

## Ethical approval and consent to participate

The study was approved by Nigeria’s National Health Research Ethics Committee (NHREC/01/01/2007-18/01/2024) and the Health Sciences Research Ethics Board at Dalhousie University (REB# 2024–7085). Informed consent was obtained from all participants before their inclusion in the study. Participants were fully informed of the study’s objectives, procedures, risks, and benefits, as well as their right to withdraw at any time without penalty. All collected data was handled with confidentiality, and no personally identifiable information will be published or shared.

## Consent for publication

Not applicable.

## Acknowledgments

This work is supported by Creating Hope in Conflict: a Humanitarian Grand Challenge; a partnership of the United States Agency for International Development (USAID), the Foreign, Commonwealth & Development Office, United Kingdom of Great Britain and Northern Ireland (FCDO), the Stabilisation and Humanitarian Aid Department, Ministry of Foreign Affairs, of the Netherlands (NL MFA), and His Majesty the King in right of Canada (“His Majesty”) represented by the Minister for International Development of Global Affairs Canada acting through The Department of Foreign Affairs, Trade and Development of Global Affairs Canada (“DFATD”) with support from Grand Challenges Canada [grant number R-HGC-POC-2408-67370]; and the Government of Canada, Canadian Institutes of Health Research, Institute of Population and Public Health [grant number PAA-192178].

## Acknowledgments

This work was supported by Grand Challenges Canada [grant number R-HGC-POC-2408-67370] and the Government of Canada, Canadian Institutes of Health Research, Institute of Population and Public Health [grant number PAA-192178].

## Data availability

All data supporting the article will be available upon reasonable request from the first and corresponding author.

## CRediT authorship contribution statement

Conceptualization: AEED, EE; Data curation: AI, AA, AEED; Formal analysis: AEED; Funding acquisition: SY, VA, COJ, RO, EE; Investigation: AEED, SY, VA, COJ, RO, EE; Methodology: AEED; Project administration: COJ, EE; Resources: EE; Supervision: RO, EE; Validation: AI, AA, AEED, COJ, RO, EE; Visualization: AEED; Writing – original draft: AEED; Writing – review & editing: AEED, II, AH, BH, COJ, RO, EE

## Declaration of competing interest

The authors declare that they have no known competing financial interests or personal relationships that could have influenced the work reported in this paper.

**Appendix A. Supplementary data Supplementary data to this article can be found online at:**

