## Supplemental File for "Mobile Life Skills Education Adoption Among Internally Displaced Persons in Northern Nigeria and Health Systems Implications for Equitable Mental Health Support: A cross-sectional study"

**Supplementary appendix for: Determinants of Interest in Mobile-Based Life Skills Education Among Internally Displaced Persons in Nigeria: Implications for Equitable Digital Mental Health Service Delivery in Fragile Settings**

**TABLE OF CONTENTS**

Study population and location………………………………………………………………………………………..4

Measures: Exposures…………………………………………………………………………………………………5

Measures: Covariates…………………………………………………………………………………………………5

Sample size calculation……………………………………………………………………………………………….6

Analysis…………………………………………………………………………………………………......................7

Objective 1. Elastic net regression…………………………………………………………………………………….7

Objective 2. Robust (modified) Poisson regression……………………………………………………………….......7

Objective 3. Sensitivity analysis for unmeasured confounding………………………………………………………8

Appendix Tables………………………………………………………………………………………………...........9

Appendix Table 1: Unadjusted Prevalence Ratios for interest in mLSE……………………………………………..9

Appendix Table 2: Conditional effects of selected IDP age groups, gender, education, and psychological counseling history on interest in mLSE……………………………………………………………………………......................10

Appendix Table 3. Sensitivity Analysis for unmeasured confounding………………………………………............11

Appendix Figures……………………………………………………………………………………………..............12

Appendix Figure 1: Predicted Probabilities of Interest in mLSE: Age and Psychological Counseling History………………………………………………………………………………………………………………..12

Appendix Figure 2: Predicted Probabilities of Interest in mLSE: Gender and Psychological Counseling History………………………………………………………………………………………………………………13

Appendix Figure 3. Predicted Probabilities of Interest in mLSE: Education and Psychological Counseling History……………………………………………………………………………………………………………….14

Appendix Figure 4. Predicted Probabilities of Interest in mLSE: Age, Gender, and Psychological Counseling History……………………………………………………………………………………………………………….15

Appendix Table 4: STROBE CHECKLIST..………………………………………………………………………..16

References…………………………………………………………………………………………………………....21

**Study population and locations**

This study analyzed cross-sectional data collected by our research team from the baseline household survey of the Rebuilding Emotional Stability and Strength Through Therapeutic and Life-Skills Education for Internally Displaced Persons in Nigeria (RESETTLE-IDPs) project in Nigeria.^1^ The RESETTLE-IDP project aims to strengthen the psychosocial wellbeing, coping abilities, and integration skills of IDPs affected by trauma and displacement to better access existing humanitarian services and sustainably integrate into host communities in Northern Nigeria. Data analysis was conducted between September 2024 and January 2025. The study questionnaire consisted of 48 questions, including sociodemographic characteristics, smartphone and mobile phone access and usage, mobile internet and social media use, and barriers to mobile access. Additional areas of focus included the potential of mobile phones for mental health and psychosocial support, mobile learning potential, and exposure to sexual violence among IDPs. Survey respondents were recruited from two IDP camps in Durumi and Wassa, Abuja, Federal Capital Territory (FCT), North Central Nigeria. Although these sites are sometimes called "settlements" due to their lack of formal infrastructure, both terms "camps" and "settlements" are often used interchangeably in humanitarian contexts.^2^ This study uses both terms for clarity. To minimize bias, participants were randomly sampled from both camps.

Abuja, FCT, is a major destination for displaced persons seeking shelter and economic opportunities after fleeing the conflict-affected states of Borno, Adamawa, and Yobe (BAY) in Northeast Nigeria.^3^ Approximately 20659 IDPs from these states live in 18 camp-like settlements across the six local government areas in Abuja, with an average of 2000 individuals per settlement.^3-5^ This makes Abuja a suitable location for the study, as it allows observing IDPs from three epicenters, Borno, Adamawa, and Yobe, within a single setting.^4-5^

The Durumi and Wassa IDP settlements present distinct characteristics while serving similar displaced populations. Durumi, situated in the urban Area 1 of the Abuja Municipal Area Council, benefits from its city location while hosting residents primarily from Borno State who fled the Boko Haram insurgency.^6-9^ The camp features specific educational programs like the School Without Walls project by Life Builders Initiative. In contrast, Wassa IDP camp, located in a more rural area of the same council, faces greater infrastructural challenges with residents living in makeshift shelters.^3-,9^ While both camps rely on nonprofit organizational support for healthcare through medical outreaches, Wassa's rural setting creates additional barriers to accessing these services.^3-,9^ Both locations house diverse populations of men, women, and children, with similar housing and water access challenges.^3-9^ However, Wassa's remote location amplifies these difficulties compared to the more centrally located Durumi camp.^3-11^

**Measures**

**Exposures**

Two key variables were used to assess the participants' experience with psychological counseling. First, participants were asked, “Have you received any psychological counseling since you were displaced?” with possible responses of “Yes,” “No,” or “I can’t remember.” This variable was recoded into a binary format, where “Yes” responses remained as “Yes,” and “No” and “I can’t remember” responses were combined into “No.” The second variable, Counseling Location, asked participants who reported receiving counseling where the counseling took place. The response options included “IDP camp,” “In the community,” “At the mosque/church,” “At the health facility,” or “Other,” and this variable was retained as categorical to explore differences in counseling experiences based on location.

The duration of stay in the IDP camp was categorized as either less than or equal to five years or six years or above.

**Covariates**

Sociodemographic factors, including the participant’s age, gender, and state of residence before displacement, were assessed as covariates. The participant’s spoken languages, including English, Hausa, Fulfulde, Igbo, Yoruba, Pidgin English, and other languages, were assessed using multiple response options. Similarly, the primary reading language was recorded with the same response options as spoken languages.

Participants’ education level was categorized into the following groups: No formal education, Islamic school, Primary school, Secondary school, University or technical college, Postgraduate degree, and Other. Employment status was assessed as a binary variable, with responses indicating whether the participant was employed. The name of the IDP settlement was also recorded as a binary variable, with responses indicating either Durumi or Wassa.

Social media usage was measured using several variables. Social usage and process usage were assessed via yes/no questions. For preferred mobile technology for support, participants were asked to indicate their preference among the following options: phone calls, Short Message Service (SMS), messaging apps, customized mobile apps, or none. Participants’ mobile internet and social media usage were also recorded. Internet access on mobile phones was a binary variable, while mobile phone use was categorized into Calls and Miscellaneous use. Participants were asked to report which social media platforms they use, e.g., WhatsApp, Facebook, Telegram, Instagram, TikTok, Snapchat, Twitter, Other, or None. The purpose of social media usage was assessed using multiple-response options, such as communication, news access, entertainment, education, community networking, business/trade, and other purposes. Social media group membership was recorded as a binary variable (Yes/No). Finally, participants were asked about their preferred content format in social media groups, including text, image, video, audio, or other formats.

To capture obstacles to mobile access, participants were asked to identify challenges they face when using mobile technology. Responses were provided in multiple categories, including “Lack of personal phone,” “Limited access to shared phones,” “Affordability issues,” “Poor connectivity,” “Lack of electricity,” “Lack of knowledge,” “Privacy/safety concerns,” “No barriers,” and “Other.” These responses were recoded into three categorical groups: Affordability/Access, Technological, and None.

**Sample size calculation**

The initial sample size calculation targeted 102 participants, based on a 60% prevalence rate of mental health and psychosocial support needs, 10% precision level, 5% error risk, and 10% non-response rate.^12-13^ To ensure sufficient statistical power (>80%) for detecting medium effect sizes (d = 0.5) and to accommodate subgroup analyses, the target sample size was increased. With a 100% response rate achieved (n = 220), the study design exceeded its initial target and provided robust power (alpha = 0.05) for analyzing primary and secondary outcomes and subgroup effects.

**Analysis**

**Objective 1: Elastic net regression**

Elastic net regression was utilized for variable selection,^14^ identifying 22 important predictors of interest in mobile-based life skills education (mLSE). A 10-fold cross-validation approach was used to optimize model parameters, and the final model, with an α value of 0.50 (balancing L1/L2 penalties), retained variables with standardized coefficients > 0.1. The performance of the final elastic net model was evaluated using the Area Under the Receiver Operating Characteristic Curve (AUC ROC), which reached 0.89 (Figure 1). Variable importance plots and a correlation matrix are presented in Figures 2 and 3.

The elastic net analysis highlighted a subset of key variables, including lack of a social media platform, being 35 years or older, duration of stay in an IDP camp, and secondary education or higher as highly influential variables. Other key variables, e.g., technological barriers, younger age (20-24 years), language spoken, and IDP camp location, were considered important but deemed less influential (Figure 2).

**Objective 2: Robust (modified) Poisson regression**

Following variable selection, differences between IDPs based on interest in mLSE were statistically assessed across the categories of each variable using χ^2^ tests (Table 1). In addition, we utilized the identified predictors in a modified robust Poisson regression model,^15^ including some other key variables., e.g., psychological counseling history and gender, that were not included in the final elastic net model but aligned with our study objectives. The binary outcome (interest in mLSE: yes/no) was modeled in the robust Poisson regression,^15^ which provided prevalence ratios (PRs) for the predictors (Table 2).^16^

This modeling approach can estimate coefficients while accounting for non-convergence, overdispersion, and heteroskedasticity in data.^17^ To ensure the robustness of our results, we calculated robust standard errors using the HC0 estimator.^18^ This step was crucial in addressing potential violations of model assumptions. The model coefficients were then exponentiated to obtain prevalence and adjusted prevalence ratios (PRs/APRs), providing a more intuitive interpretation of the results (Table 2, Figure 4, Appendix Table 1). We computed 95% confidence intervals for these effect measures using robust standard errors. *P*-values less than .05 (2-tailed) were considered statistically significant. For each predictor in the robust Poisson regression model, the prevalence ratio, *p_xi_* /*p_xj_*, was interpreted as the change in event probability when transitioning from level *j* of the predictor to level *i*.^19-20^ When the variable whose prevalence ratio is to be estimated interacted with another variable in the model, the prevalence ratio varied based on the interacting variable(s) levels.^19-20^

To evaluate the predictive accuracy of significant interaction effects on IDP interest in mLSE (i.e., number of events), marginal standardized plots^21-23^ were generated to depict the probabilities for key interactions: age and psychological counseling history, gender and psychological counseling history, education and psychological counseling history, and the combined effects of age, gender, and psychological counseling history (Appendix Figures 1-4). Marginal standardization is a suitable method for inferring population-level effects.^21-23^

By utilizing this multistage analytical process, we were able to generate reliable estimates of the factors influencing interest in mLSE among IDPs while simultaneously controlling for potential confounders (Appendix Table 2).

**Objective 3: Sensitivity analysis for unmeasured confounding**

E-values were used for sensitivity analysis to address unmeasured confounding (Appendix Table 3). Sensitivity analysis showed relatively modest E-values (Appendix Table 3). Although the associations observed in this study appear robust to a substantial degree of potential unmeasured confounding, especially when interpreted in the context of similar studies^24-25^, it is plausible that unaccounted residual or unmeasured confounders not captured in the current analysis could affect some associations. In short, a small effect size from such unmeasured confounders might be sufficient to explain some of our reported associations.^24-25^

**Appendix Tables**

**Appendix Table 1: Unadjusted Prevalence Ratios for interest in mLSE**

| mLSE | PR | Robust  Std. Err. | z | P>\|z\| | [95% Conf. Interval] |
| --- | --- | --- | --- | --- | --- |
| AGE  13-19 years  20-24 years  25-29 years  30-34 years  35+ years | 1.00 (base)  0.75  0.68  0.39  0.24 | 0.08  0.07  0.06  0.05 | -2.63  -3.84  -5.69  -6.88 | 0.0090  <0.0001  <0.0001  <0.0001 | 0.61 0.93  0.56 0.83  0.28 0.54  0.16 0.36 |
| GENDER  Female  Male | 1.00 (base)  1.24 | 0.19 | 1.44 | 0.145 | 0.93 1.67 |
| EDUCATION  Islamic education  No formal education  Primary Education  Secondary Education or Higher | 1.00 (base)  0.37  1.26  2.45 | 0.12  0.29  0.40 | -3.08  1.01  5.46 | 0.0020  0.31  <0.0001 | 0.20 0.70  0.81 1.96  1.78 3.39 |
| EMPLOYMENT  No  Yes | 1.00 (base)  1.28 | 0.18 | 1.73 | 0.084 | 0.97 1.69 |
| BARRIER  Accessibility/Affordability  None  Technological | 1.00 (base)  1.47  0.83 | 0.25  0.13 | 2.28  -1.26 | 0.023  0.21 | 1.05 2.04  0.61 1.11 |
| IDP CAMP LOCATION  Durumi IDP Camp  Wassa IDP Camp | 1.00 (base)  1.23 | 0.17 | 1.47 | 0.14 | 0.93 1.61 |
| ENGLISH  No  Yes | 1.00 (base)  1.76 | 0.22 | 4.44 | <0.0001 | 1.37 2.25 |
| HAUSA  No  Yes | 1.00 (base)  0.85 | 0.13 | -1.07 | 0.29 | 0.63 1.15 |
| MOBILE PHONE USE  Calls  Miscellaneous | 1.00 (base)  2.33 | 0.29 | 6.68 | <0.0001 | 0.28 0.43 |
| NO SOCIAL MEDIA PLATFORM  False  True | 1.00 (base)  0.39 | 0.04 | -10.22 | <0.0001 | 0.32 0.47 |
| PSYCHOLOGICAL COUNSELING HISTORY  False  True | 1.00 (base)  1.51 | 0.20 | 3.04 | 0.0020 | 1.16 1.96 |
| DURATION IN CAMP  0-5 years  6+ years | 1.00 (base)  0.40 | 0.05 | -7.79 | <0.0001 | 0.32 0.50 |

| mLSE | APR | Robust  Std. Err. | z | P>\|z\| |  | [95% Conf. Interval] |
| --- | --- | --- | --- | --- | --- | --- |
| AGE*GENDER: 20-24 years*Male | 1.75 | 0.44 | 2.19 | 0.028 | 1.06 | 2.88 |
| AGE*GENDER: 30-34 years*Male | 0.09 | 0.10 | -2.22 | 0.026 | 0.01 | 0.75 |
| AGE*PSYCHOLOGICAL COUNSELING HISTORY: 20-24 years*True | 3.49 | 1.26 | 3.45 | 0.0010 | 1.72 | 7.10 |
| GENDER*PSYCHOLOGICAL COUNSELING HISTORY: Male*True | 0.33 | 0.09 | -3.95 | <0.0001 | 0.19 | 0.57 |
| EDUCATION*PSYCHOLOGICAL COUNSELING HISTORY: Primary Education*True | 2.76 | 0.86 | 3.23 | 0.0010 | 1.49 | 5.10 |

**Appendix Table 2: Conditional effects of selected IDP age groups, gender, education, and psychological counseling history on interest in mLSE**

**Appendix Table 3. Sensitivity Analysis for unmeasured confounding**

| mLSE | APR | [95% Conf. Interval] | EValue | [95% Conf. Interval] |
| --- | --- | --- | --- | --- |
| AGE    30-34 years  35+ years | 0.61  0.51 | 0.41 0.92  0.34 0.77 | 2.66  3.33 | NA 1.39  NA 1.92 |
| EDUCATION    No formal education    Secondary Education or Higher | 0.50  2.27 | 0.27 0.94    1.59 3.26 | 3.41  3.97 | NA 1.32  2.56 NA |
| BARRIER    None | 1.57 | 1.07 2.30 | 2.52 | 1.34 NA |
| IDP CAMP LOCATION    Wassa IDP Camp | 1.64 | 1.21 2.23 | 2.67 | 1.71 NA |
| NO SOCIAL MEDIA PLATFORM    True | 0.69 | 0.53 0.89 | 2.26 | NA 1.50 |
| DURATION IN CAMP    6+ years | 0.65 | 0.49 0.86 | 2.45 | NA 1.60 |

NA: Not Applicable

**Appendix Figures**

**Appendix Figure 1: Predicted Probabilities of Interest in mLSE: Age and Psychological Counseling History**

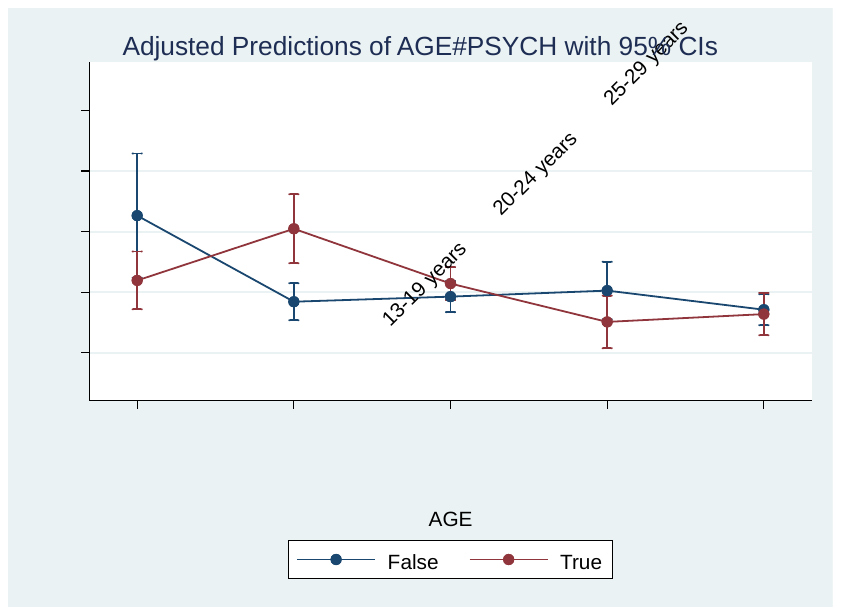

**Appendix Figure 2: Predicted Probabilities of Interest in mLSE: Gender and Psychological Counseling History**

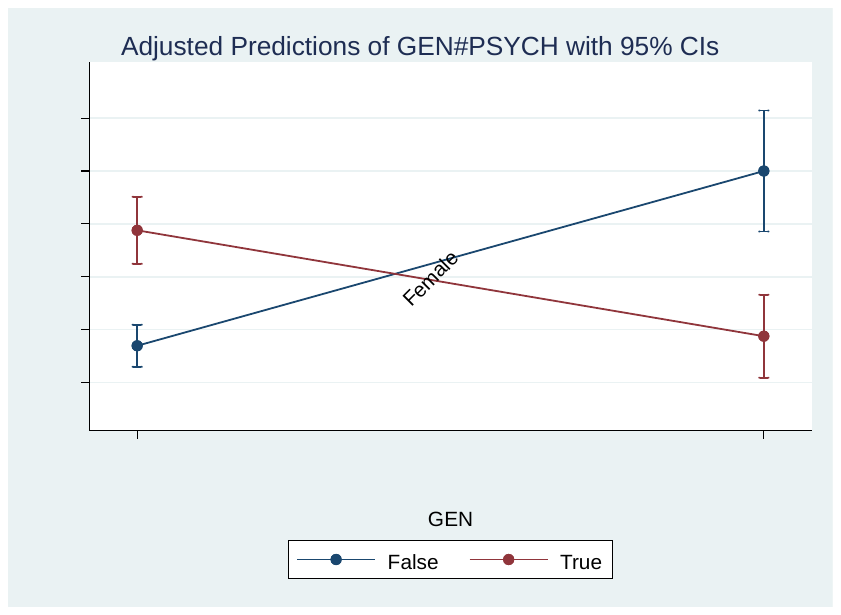

**Appendix Figure** **3. Predicted Probabilities of Interest in mLSE: Education and Psychological Counseling History**

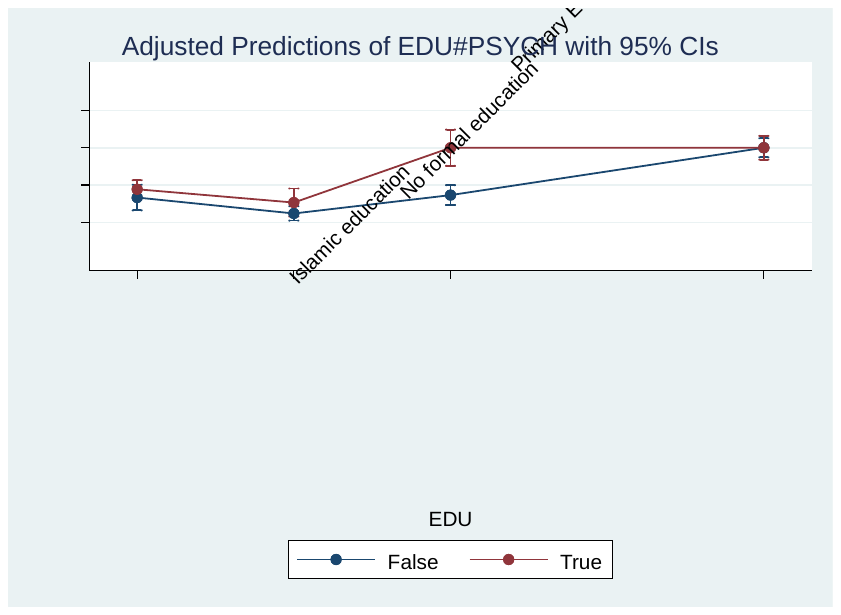

**Appendix Figure 4. Predicted Probabilities of Interest in mLSE: Age, Gender, and Psychological Counseling History**

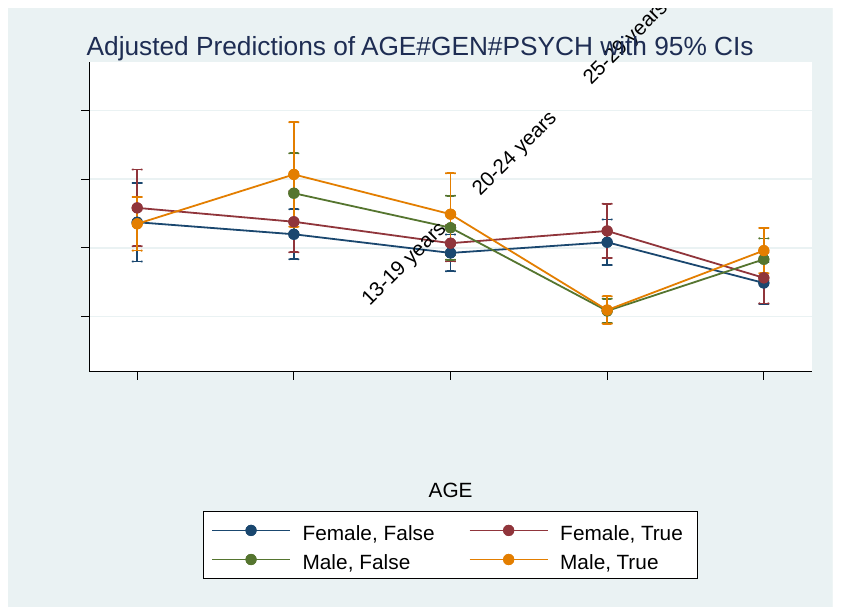

Appendix Table 4: STROBE CHECKLIST

STROBE Statement—Checklist of items that should be included in reports of ***cross-sectional studies***

|  | **Item No** | **Recommendation** | **Page No** |
| --- | --- | --- | --- |
| **Title and abstract** | 1 | (a) Indicate the study’s design with a commonly used term in the title or the abstract   A cross-sectional study | 1 |
|  |  | (*b*) Provide in the abstract an informative and balanced summary of what was done and what was found | 2 |
| **Introduction** | | | |
| Background/rationale | 2 | Explain the scientific background and rationale for the investigation being reported | 1-4 |
| Objectives | 3 | State specific objectives, including any prespecified hypotheses | 1-4 |
| **Methods** | | | |
| Study design | 4 | Present key elements of study design early in the paper | 4-5 |
| Setting | 5 | Describe the setting, locations, and relevant dates, including periods of recruitment, exposure, follow-up, and data collection | 4-5,  Appendix p.4 |
| Participants | 6 | (*a*) Give the eligibility criteria, and the sources and methods of selection of participants | 4-5 Appendix p.4 |
| Variables | 7 | Clearly define all outcomes, exposures, predictors, potential confounders, and effect modifiers. Give diagnostic criteria, if applicable | 5  Appendix pp. 3-4 |
| Data sources/ measurement | 8* | For each variable of interest, give sources of data and details of methods of assessment (measurement). Describe comparability of assessment methods if there is more than one group  Appendix Table 3 | 4-5 |
| Bias | 9 | Describe any efforts to address potential sources of bias  Appendix Table 3 | 9, 12 |
| Study size | 10 | Explain how the study size was arrived at | Appendix p.6 |
| Quantitative variables | 11 | Explain how quantitative variables were handled in the analyses. If applicable, describe which groupings were chosen and why | 5, Appendix p.6 |
| Statistical methods | 12 | (*a*) Describe all statistical methods, including those used to control for confounding | 12, Appendix p.8, p.11 |
|  |  | (*b*) Describe any methods used to examine subgroups and interactions | 12-15, Appendix p.8 |
|  |  | (*c*) Explain how missing data were addressed |  |
|  |  | (*d*) If applicable, describe analytical methods taking account of sampling strategy | 5-6, Appendix p.6 |
|  |  | (*e*) Describe any sensitivity analyses | 12-16, Appendix p.6 |
| **Results** | | | |
| Participants | 13* | (a) Report numbers of individuals at each stage of study—eg numbers potentially eligible, examined for eligibility, confirmed eligible, included in the study, completing follow-up, and analysed | 1, 6, 13 |
|  |  | (b) Give reasons for non-participation at each stage |  |
|  |  | (c) Consider use of a flow diagram |  |
| Descriptive data | 14* | (a) Give characteristics of study participants (eg demographic, clinical, social) and information on exposures and potential confounders | 13, Appendix p.5 |
|  |  | (b) Indicate number of participants with missing data for each variable of interest |  |
| Outcome data | 15* | Report numbers of outcome events or summary measures | 13-15 |
| Main results | 16 | (*a*) Give unadjusted estimates and, if applicable, confounder-adjusted estimates and their precision (eg, 95% confidence interval). Make clear which confounders were adjusted for and why they were included | 9-15, Appendix pp.11-15 |
|  |  | (*b*) Report category boundaries when continuous variables were categorized |  |
|  |  | (*c*) If relevant, consider translating estimates of relative risk into absolute risk for a meaningful time period |  |
| Other analyses | 17 | Report other analyses done—eg analyses of subgroups and interactions, and sensitivity analyses | 15-16, Appendix p.5 |
| **Discussion** | | | |
| Key results | 18 | Summarise key results with reference to study objectives | 13-16 |
| Limitations | 19 | Discuss limitations of the study, taking into account sources of potential bias or imprecision. Discuss both direction and magnitude of any potential bias | 20 |
| Interpretation | 20 | Give a cautious overall interpretation of results considering objectives, limitations, multiplicity of analyses, results from similar studies, and other relevant evidence | 16-20 |
| Generalisability | 21 | Discuss the generalisability (external validity) of the study results | 20 |
| **Other information** | | | |
| Funding | 22 | Give the source of funding and the role of the funders for the present study and, if applicable, for the original study on which the present article is based | 21 |

*Give information separately for exposed and unexposed groups.

**Note:** An Explanation and Elaboration article discusses each checklist item and gives methodological background and published examples of transparent reporting. The STROBE checklist is best used in conjunction with this article (freely available on the Web sites of PLoS Medicine at http://www.plosmedicine.org/, Annals of Internal Medicine at http://www.annals.org/, and Epidemiology at http://www.epidem.com/). Information on the STROBE Initiative is available at [www.strobe-statement.org](http://www.strobe-statement.org).
